# Long-term prognosis of adults with moderate-severe SARS-CoV-2 lower respiratory tract infection managed in primary care: prospective cohort study

**DOI:** 10.1101/2022.06.07.22276108

**Authors:** T.N. Platteel, J.C. Koelmans, D. Cianci, N.J.H. Broers, E.G.P.M. de Bont, J.W.L. Cals, R.P. Venekamp, T.J.M. Verheij

**Affiliations:** Julius Center for Health Sciences and Primary Care, University Medical Center Utrecht; Department of Family Medicine, Care and Public Health Research Institute (CAPHRI), Maastricht University

**Author notes:** **Corresponding author:** T.N. Platteel, Heidelberglaan 100, 3584 CX Utrecht, The Netherlands. Email addresses. E.G.P.M. de Bont.

## Abstract

**Objectives:** To determine differences in health-related quality of life (HRQoL) and presence and duration of symptoms between adults with and without established SARS-CoV-2 moderately severe lower respiratory tract infection (LRTI) in the 12 months following their primary care visit.

**Design:** Prospective cohort study

**Setting:** 35 general practices in the provinces Noord-Brabant and Utrecht, the Netherlands.

**Participants:** Individuals aged ≥18 years who presented to their general practitioner (GP) with a moderately severe LRTI during the first COVID-19 waive in The Netherlands (March-June 2021) underwent serology testing (participants, GPs and study personnel remained blinded for serology outcomes during study conduct) and completed baseline and follow-up questionnaires. Of the 315 participants who gave consent, 277 (88%) were suitable for inclusion in the analyses. Complete follow-up date was available in 97% of participants.

**Main outcome measures:** 1) Scores of SF-36; physical component summary (PCS), mental component summary (MCS) and subscales. 2) Risk of any and individual persisting symptoms (of cough, dyspnea, chest pain, fatigue, brain fog, headache, and anosmia/ageusia) over time.

**Results:** The change in SF-36 PSC (p=0.13), MCS (p=0.30), as well as subscale scores, over time did not differ between SARS-CoV-2 serology positive and negative participants after adjusting for sex, age, BMI, diabetes and chronic pulmonary conditions. The risk of any persisting symptom over time did not significantly differ between the groups (aHR 0.61, 95% CI 0.33-1.15), nor did the risk of individual symptoms.

**Conclusions:** In the 12 months following their moderately severe LRTI, primary care patients with and without confirmed SARS-CoV-2 infection had a comparable HRQoL profile. Albeit a considerable proportion of patients reported persistent symptoms, there was no evidence of a difference in the course of symptoms over time between patients with and without confirmed SARS-CoV-2 infection.

**Trial registration:** Dutch Trial Register (NTR) number NL8729

## Introduction

Coronavirus disease 2019 (COVID-19) caused by severe acute respiratory syndrome coronavirus-2 (SARS-CoV-2) has led to a global healthcare and societal crisis. During the early stages of the pandemic, the impact of COVID-19 on hospital and ICU admissions and mortality was the main focus of policy makers and media. However, COVID-19 has a much broader impact on society beyond flooding of secondary care, with a substantial proportion of non-hospitalised COVID-19 patients suffering from persistent symptoms such as fatigue, dyspnea, chest pain, anosmia, mental health problems, and cognitive deficits (1, 2). This condition in which symptoms persist beyond 4 weeks has been described as post-acute COVID-19 or long-COVID (1).

Although evidence on long-COVID is accumulating rapidly, information about its incidence and impact in patients treated in primary care is scarce. The few studies available reported long-COVID symptoms in 28-64% of patients at 4 weeks to 12 months after symptom onset (3-8). These figures are, however, difficult to interpret due to the absence of an adequate control group. This is an important omission because lower respiratory tract infections (LRTIs) caused by other viral and bacterial pathogens can also lead to long-lasting sequelae. In addition, perceived symptoms might be influenced by the negative impact of generic COVID-19 societal restrictions on wellbeing and patients’ awareness of having this new viral infection with unknown course of disease (9).

A few studies tried to mitigate this risk of bias by including a SARS-CoV-2 negative control group (10-14). However, in order to truly prevent bias caused by subjective outcome assessment, a double-blinded study design is warranted. The lack of SARS-CoV-2 test availability for patients presenting to primary care with LRTIs during the first wave of the COVID-19 pandemic (March-May 2020) in The Netherlands provided us with the unique opportunity to conduct such a study. To this end, we prospectively followed patients presenting to primary care with a moderately severe LRTI for 12 months and determined the difference in health-related quality of life (HRQoL) and presence and duration of long-COVID symptoms between those with and without established SARS-CoV-2 infection based on serology testing while keeping participants, General Practitioners (GPs) and study personnel blinded for serology outcomes during the study conduct.

## Methods

### Setting and study population

Between March 1^st^ and June 1^st^ 2020, adult patients presenting to primary care with a moderately severe LRTI were retrospectively selected from 35 GP practices in the provinces of Noord-Brabant and Utrecht, the Netherlands. Moderately severe LRTI was defined as International Classification of Primary Care (ICPC) code R81 pneumonia or either code R02 (dyspnea), R05 (cough), R74 (respiratory tract infection), R78 (bronchitis) or R83 (other respiratory tract infection), and in which the GP decided to treat LRTI symptoms with an oral antibiotic. At the time of the study no widescale COVID-19 PCR tests were available in The Netherlands. Only hospitalized patients and health care workers were tested. Patients who were admitted to the hospital in the 14 days before or after the index consultation (i.e. patients’ first primary care contact for the LRTI episode), those with a life expectancy of less than a year and those who were not capable of completing online, paper or telephone questionnaires, according to their GP, were excluded from participation.

Potentially eligible participants were informed about the study by their GP. Those who expressed interest in study participation, received verbal and written information from the study team. Full written informed consent was then obtained from those who agreed to participate.

### Study procedures, baseline and follow-up measurements

At baseline, defined as moment of inclusion in the study, a venous blood sample was taken from participants for Abbott Alinity SARS-CoV-2 spike-specific quantitative IgG serology testing, a highly specific serology test with IgG antibodies remaining detectable up to seven months after symptom onset in at least 92% of patients (13). Participants, GPs and study personnel remained blinded for the serology test results until follow-up and analyses were completed.

Next, participants were asked to complete a questionnaire about demographics, presence and severity of symptoms (cough, dyspnea, chest pain, fatigue, brain fog, headache, anosmia/ageusia using a 0-4 Likert scale (0: absent, 1: a little, 2: moderate, 3: much and 4: very much) at time of the index consultation (recall) and at baseline, and their HRQoL two weeks prior to the index consultation (recall) and at baseline using the SF-36 questionnaire (15). This validated HRQoL questionnaire consists of a single item of health transition (HT) and a further 35 items which can be divided into 8 subscales: (1) physical function (PF), (2) limitations due to physical health problems (role physical, RP), (3) bodily pain (BP), (4) general health (GH), (5) vitality (VT), (6) social functioning (SF), (7) limitations due to emotional health problems (role-emotional, RE), and (8) mental health (MH). The scores of SF-36 between 0 and 100 were assigned to each domain, with higher scores indicating more favourable functional status. The eight subdomain scores were aggregated into two summary measures: physical component summary (PCS) scores and mental component summary (MCS) scores.

Follow-up HRQoL and symptom questionnaires were performed every 3 months for a total of 12 months after the index consult. In addition, participants were asked to report any positive SARS-COV-2 test result during follow-up.

Finally, data on participants’ signs and symptoms at the index consultation, comorbidities and medication use, as well as results of SARS-CoV-2 reverse-transcriptase polymerase chain reaction (RT-PCR) and serology were extracted from patients’ electronic primary care health records.

### Outcome measures

The primary outcome of interest was change in participants’ SF-36 PCS scores over time. Secondary outcomes included changes over time in SF-36 MCS scores and the eight individual SF-36 subscales, as well as the time to resolution of symptoms (cough, dyspnea, chest pain, fatigue, brain fog, headache, anosmia/ageusia).

### Patient involvement

Patients were not involved in the design of the study.

### Sample size considerations

We assumed the mean SF-36 PCS score to be 50 with a standard deviation (SD) of 10 (16). Assuming that at least 20% of moderately severe LRTI were caused by SARS-CoV-2, an effective sample size of 274 patients would therefore be required to achieve 80% power to detect a clinically significant difference of three or more points in the SF-36 PCS score between groups (17).

### Statistical analyses

Baseline characteristics were compared between those with and without established SARS-CoV-2 infection (based on serology testing) using appropriate statistical tests.

For the primary analysis, we used linear mixed effects model with SF-36 PSC scores as dependent variable and SARS-CoV-2 serology test result, time, age, gender, body mass index (BMI), diabetes, chronic pulmonary diseases and the interaction term between time and SARS-CoV-2 serology test result as fixed effect. The random intercept and slope for time account for repeated measurements. The difference in change of SF-36 PSC score from 2 weeks prior to 12 months after the index consultation between the two groups (SARS-CoV-2 serology positive versus negative) were estimated from the model and presented with accompanying 95% confidence intervals (CIs).

We estimated the difference in change of SF-36 MSC scores and the eight individual SF-36 subscales between groups using a similar approach.

For time to resolution of symptoms, we performed Kaplan-Meier survival analyses with log rank tests. Patients with persisting symptoms were censored at the end of follow-up. Unadjusted hazard ratios (HRs) with accompanying 95% CIs were calculated using Cox proportional hazard modelling. Age, gender, and comorbidities (chronic pulmonary disease, cardiovascular disease, and diabetes) were added to the models as potential confounders to derive adjusted HRs with 95% CIs. For each covariate, the proportional hazards assumption was tested using the supremum test; when it was clearly violated, we added time-dependent coefficients for those covariates (18). In addition, we assessed whether chronic pulmonary disease, cardiovascular disease and diabetes modified the effect by adding them as interaction term with SARS-CoV-2 serology test result to the model (with P<0.05 indicating significant effect modification).

For the analyses several assumptions were made. Primarily, the study SARS-CoV-2 serology test result was used to determine whether the moderately severe LRTI at index consultation was attributed to SARS-CoV-2 infection. However, if there were discrepancies between the serology test result and any routine care RT-PCR- or serology test result performed prior to the study serology test, the participant was excluded from the analysis. Participants who tested RT-PCR positive between one month after the index consultation and 14 days before the study serology were considered SARS-CoV-2 negative until the day of the positive RT-PCR result. Participants with SARS-CoV-2 negative serology test who experienced a SARS-CoV-2 infection during follow-up were censored from the moment of positive SARS-CoV-2 test onwards.

Several post-hoc sensitivity analyses were performed. First, we excluded all participants who knew their SARS-CoV-2 status, and thus were “deblinded”, prior to completing the first questionnaire. If participants were deblinded after completion of the first questionnaire, they were censored from the moment of deblinding. In a second sensitivity analysis, we excluded all participants with chronic pulmonary disease, as patients with chronic symptoms, present before the index consultation, could lead to an overestimation of persisting symptoms in the SAR-CoV-2 serology negative group. Finally, in a sensitivity analysis of the duration of symptoms, we defined a Likert scale symptom score ≤2 as symptom resolution.

Data were analyzed using IBM SPSS Statistics version 26.0.0.1 for windows. SAS version 9.4 was used for the linear mixed effects model and the proportional hazards checks. Statistical significance was assumed at a p-value <0.05.

## Results

### Study population

Of the 443 patients with moderately severe LRTI that were approached for participation, 315 consented to participate. Of these, 277 (88%) were suitable for inclusion in the analyses. We obtained complete follow-up data in 97% of the 277 patients (Figure 1).

**Figure 1.**
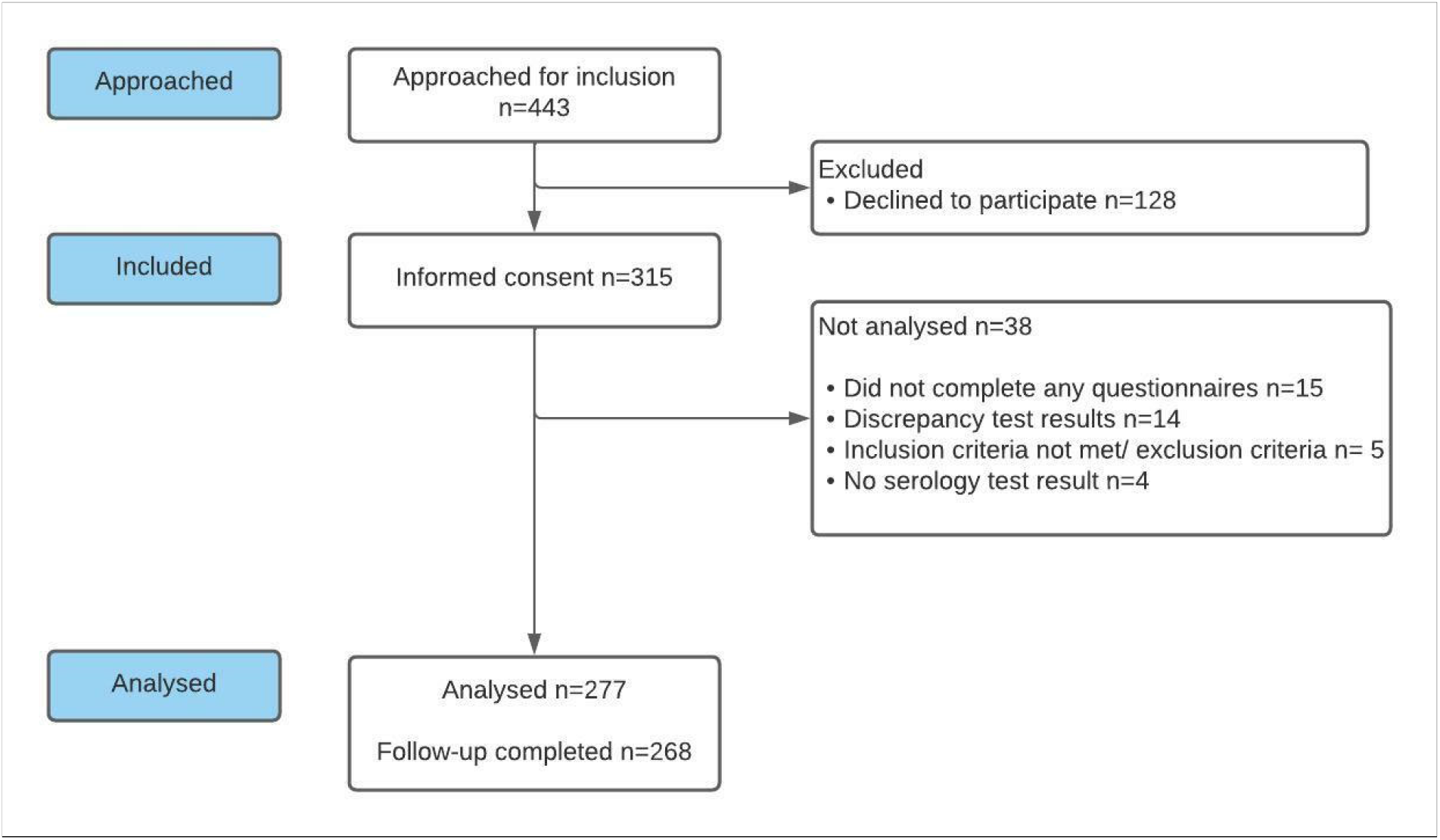
Patient selection flowchart

The first follow-up questionnaire was completed after a median of 7.8 months (IQR 7.1-8.4) after the index consultation. Serology testing was performed after a median of 7.6 months (IQR 6.9-8.4).

### Baseline characteristics

The SARS-CoV-2 serology test was positive in 19.9% (55/277) of participants (Table 1). Participants with a positive serology test were older (mean age 59.9 years [SD 10.7] vs 54.8 years [SD 14.6]) and were more likely to have diabetes (20.0% vs 7.2%). Active smoking (17.1% vs 1.8%) and chronic pulmonary conditions (27.0% vs 10.9%) were more prevalent in SARS-CoV-2 serology negative participants.

**Table 1.**
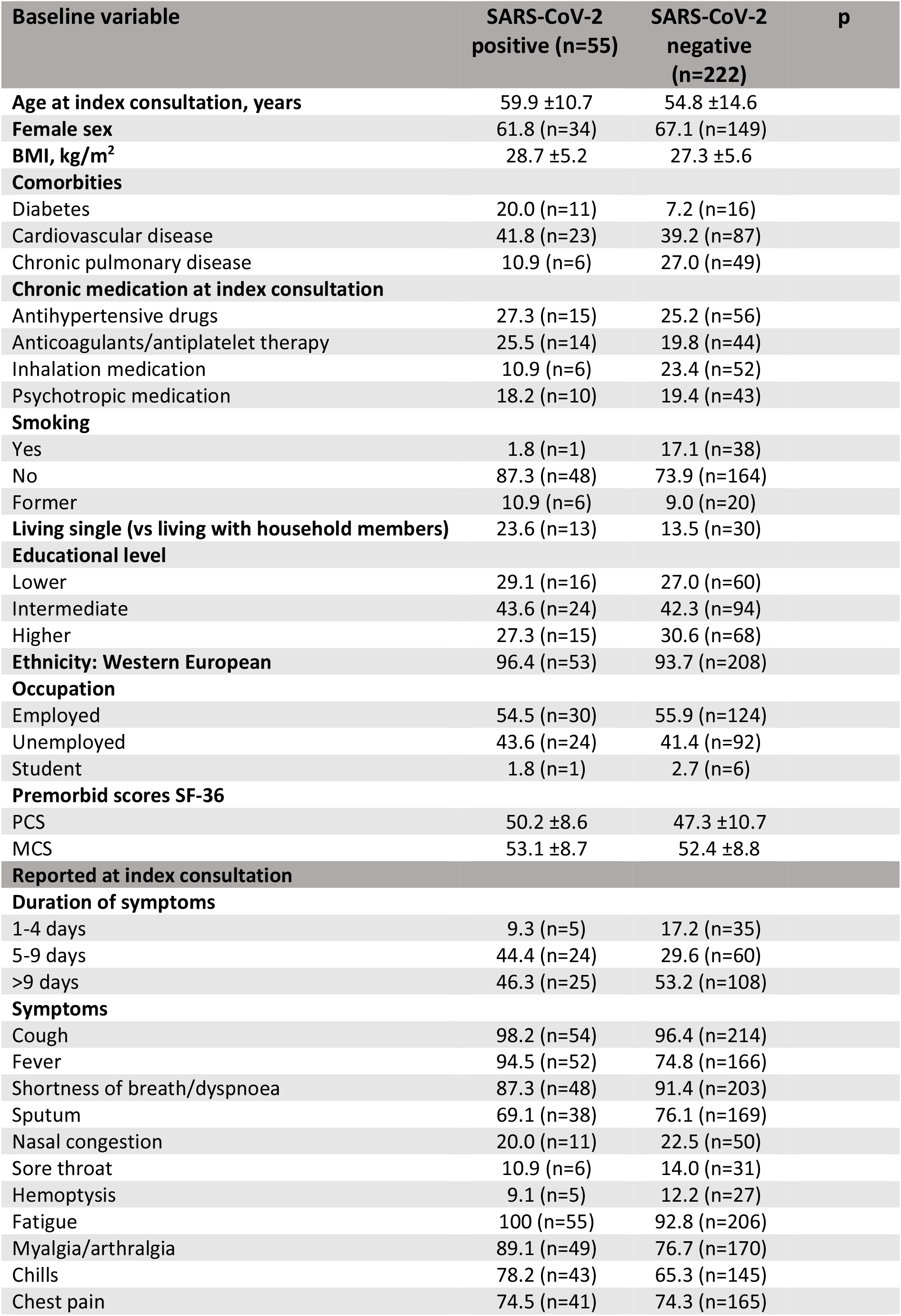
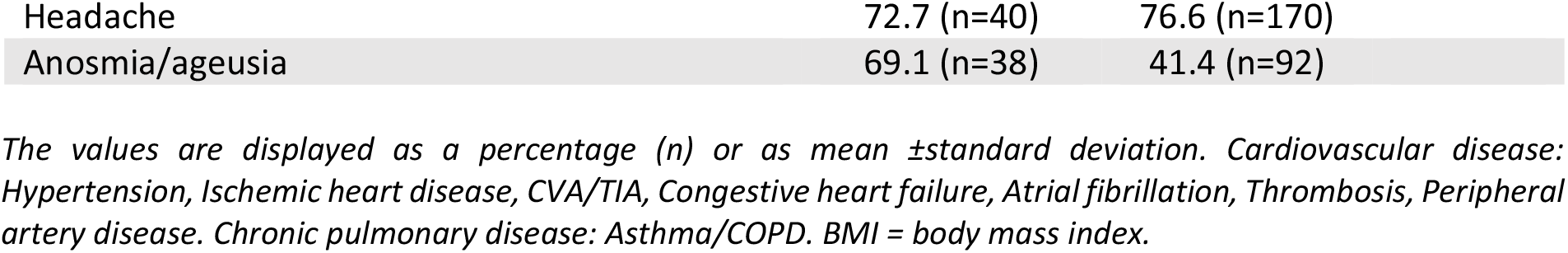
Baseline characteristics, including demographics and clinical presentation of SARS-CoV-2 positive and negative participants.

| Baseline variable | SARS-CoV-2 positive (n=55) | SARS-CoV-2 negative (n=222) | p |
| --- | --- | --- | --- |
| **Age at index consultation, years** | 59.9 ±10.7 | 54.8 ±14.6 |  |
| **Female sex** | 61.8 (n=34) | 67.1 (n=149) |  |
| **BMI, kg/m<sup>2</sup>** | 28.7 ±5.2 | 27.3 ±5.6 |  |
| **Comorbidities** |  |  |  |
| Diabetes | 20.0 (n=11) | 7.2 (n=16) |  |
| Cardiovascular disease | 41.8 (n=23) | 39.2 (n=87) |  |
| Chronic pulmonary disease | 10.9 (n=6) | 27.0 (n=49) |  |
| **Chronic medication at index consultation** |  |  |  |
| Antihypertensive drugs | 27.3 (n=15) | 25.2 (n=56) |  |
| Anticoagulants/antiplatelet therapy | 25.5 (n=14) | 19.8 (n=44) |  |
| Inhalation medication | 10.9 (n=6) | 23.4 (n=52) |  |
| Psychotropic medication | 18.2 (n=10) | 19.4 (n=43) |  |
| **Smoking** |  |  |  |
| Yes | 1.8 (n=1) | 17.1 (n=38) |  |
| No | 87.3 (n=48) | 73.9 (n=164) |  |
| Former | 10.9 (n=6) | 9.0 (n=20) |  |
| **Living single (vs living with household members)** | 23.6 (n=13) | 13.5 (n=30) |  |
| **Educational level** |  |  |  |
| Lower | 29.1 (n=16) | 27.0 (n=60) |  |
| Intermediate | 43.6 (n=24) | 42.3 (n=94) |  |
| Higher | 27.3 (n=15) | 30.6 (n=68) |  |
| **Ethnicity: Western European** | 96.4 (n=53) | 93.7 (n=208) |  |
| **Occupation** |  |  |  |
| Employed | 54.5 (n=30) | 55.9 (n=124) |  |
| Unemployed | 43.6 (n=24) | 41.4 (n=92) |  |
| Student | 1.8 (n=1) | 2.7 (n=6) |  |
| **Premorbid scores SF-36** |  |  |  |
| PCS | 50.2 ±8.6 | 47.3 ±10.7 |  |
| MCS | 53.1 ±8.7 | 52.4 ±8.8 |  |
| **Reported at index consultation** |  |  |  |
| **Duration of symptoms** |  |  |  |
| 1-4 days | 9.3 (n=5) | 17.2 (n=35) |  |
| 5-9 days | 44.4 (n=24) | 29.6 (n=60) |  |
| >9 days | 46.3 (n=25) | 53.2 (n=108) |  |
| **Symptoms** |  |  |  |
| Cough | 98.2 (n=54) | 96.4 (n=214) |  |
| Fever | 94.5 (n=52) | 74.8 (n=166) |  |
| Shortness of breath/dyspnoea | 87.3 (n=48) | 91.4 (n=203) |  |
| Sputum | 69.1 (n=38) | 76.1 (n=169) |  |
| Nasal congestion | 20.0 (n=11) | 22.5 (n=50) |  |
| Sore throat | 10.9 (n=6) | 14.0 (n=31) |  |
| Hemoptysis | 9.1 (n=5) | 12.2 (n=27) |  |
| Fatigue | 100 (n=55) | 92.8 (n=206) |  |
| Myalgia/arthralgia | 89.1 (n=49) | 76.7 (n=170) |  |
| Chills | 78.2 (n=43) | 65.3 (n=145) |  |
| Chest pain | 74.5 (n=41) | 74.3 (n=165) |  |

|  |  |  |
| --- | --- | --- |
| Headache | 72.7 (n=40) | 76.6 (n=170) |
| Anosmia/ageusia | 69.1 (n=38) | 41.4 (n=92) |

*The values are displayed as a percentage (n) or as mean ± standard deviation. Cardiovascular disease: Hypertension, Ischemic heart disease, CVA/TIA, Congestive heart failure, Atrial fibrillation, Thrombosis, Peripheral artery disease. Chronic pulmonary disease: Asthma/COPD. BMI = body mass index.*

At baseline there were no significant differences in the SF-36 PCS and MCS scores between SARS-CoV-2 serology negative and positive participants (50.2 [SD 8.6] vs 47.3 [SD 10.7] and 53.1 [SD 8.7] vs 52.4 [SD8.8], respectively). Symptoms more prevalent in SARS-CoV-2 positive participants at index consultation were fever (94.5% vs 74.8%), myalgia/arthralgia (89.1% vs 76.6%), fatigue (100% vs 92.8%), and anosmia/ageusia (69.1% vs 41.4%).

### HRQoL scores over time

The change in SF-36 PSC score over time did not differ between SARS-CoV-2 serology positive and negative participants (p=0.127) after adjusting for sex, age, BMI, diabetes and chronic pulmonary conditions (Figure 2).

**Figure 2.**
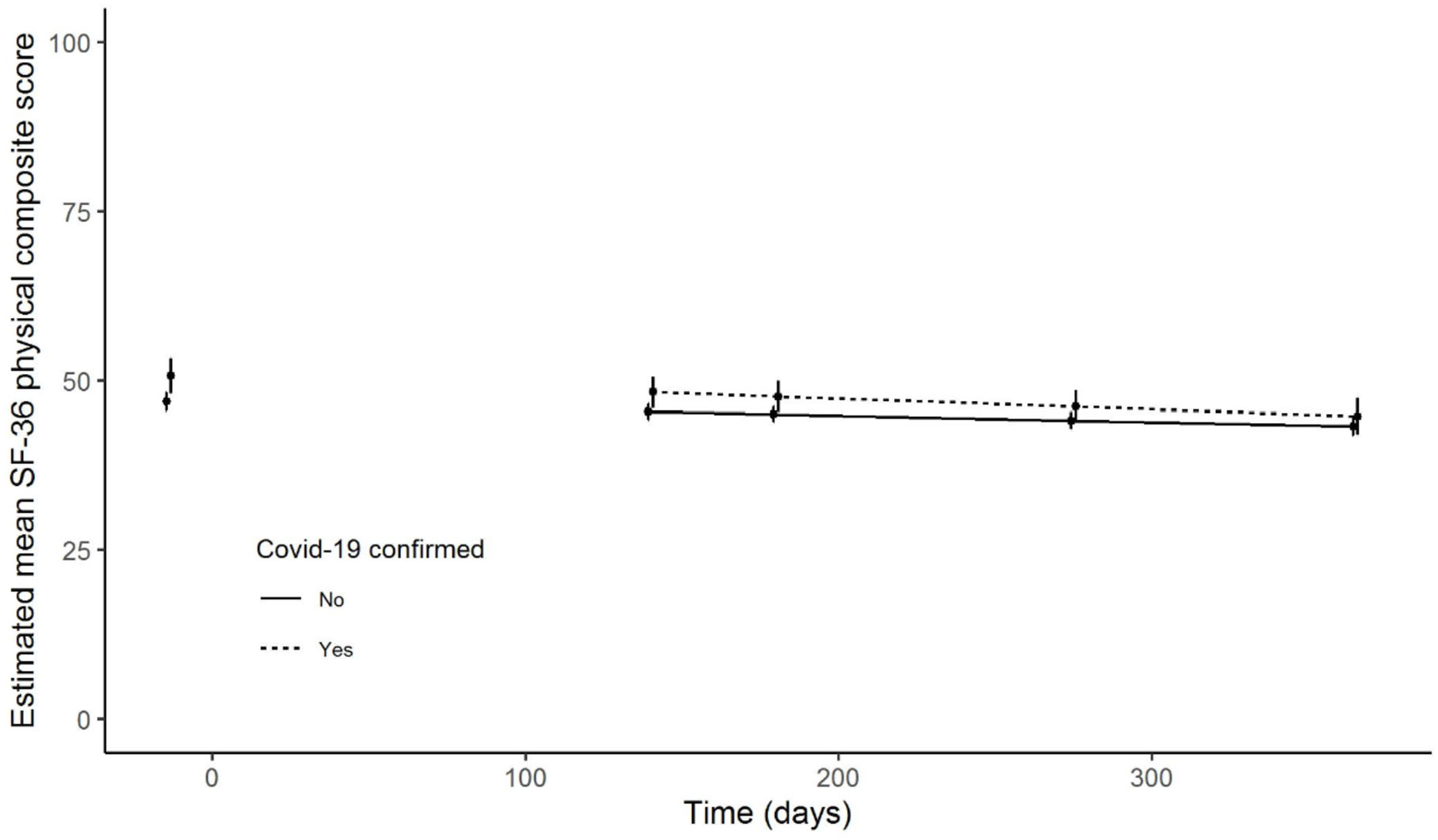
Estimated mean Physical Component Summary (PCS) score (SF-36) over time, based on mixed model.

Similarly, the change in SF-36 MCS score over time (p=0.302), did not differ between SARS-CoV-2 serology positive and negative participants (Appendix S1). The same accounts for the change in the individual eight SF-36 subscales (data not shown). Sensitivity analyses in which participants were censored from the moment of deblinding yielded similar results.

### Time to resolution of symptoms

At least one persisting symptom 4 weeks after the index consultation was reported in 80% of SARS-CoV-2 serology positive participants and in 83.8% of SARS-CoV-2 serology negative participants. Dyspnea (serology positive vs negative group: 54.5% vs 58.1%), fatigue (63.6% vs 68.0%), cough (45.5% vs 46.8%) and brain fog (43.6% vs 37.8%) were the most commonly reported persisting symptoms (Table 2).

**Table 2.**
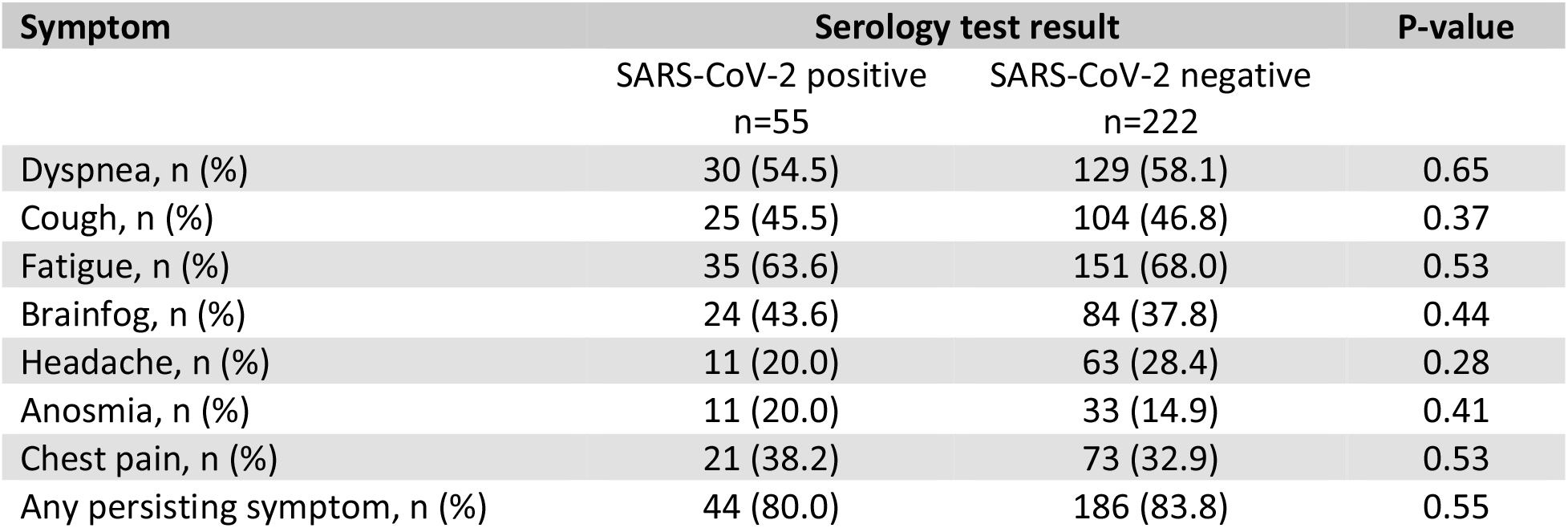
Persisting symptoms 4 weeks after the index consultation

| Symptom | Serology test result |  | P-value |
| --- | --- | --- | --- |
|  | SARS-CoV-2 positive<br>n=55 | SARS-CoV-2 negative<br>n=222 |  |
| Dyspnea, n (%) | 30 (54.5) | 129 (58.1) | 0.65 |
| Cough, n (%) | 25 (45.5) | 104 (46.8) | 0.37 |
| Fatigue, n (%) | 35 (63.6) | 151 (68.0) | 0.53 |
| Brainfog, n (%) | 24 (43.6) | 84 (37.8) | 0.44 |
| Headache, n (%) | 11 (20.0) | 63 (28.4) | 0.28 |
| Anosmia, n (%) | 11 (20.0) | 33 (14.9) | 0.41 |
| Chest pain, n (%) | 21 (38.2) | 73 (32.9) | 0.53 |
| Any persisting symptom, n (%) | 44 (80.0) | 186 (83.8) | 0.55 |

The Kaplan Meier survival curves (12-month follow-up) for any persisting symptom were comparable between SARS-CoV-2 serology negative and positive participants and log rank tests showed no significant differences (Figure 3, Table 3). The same accounts for the Kaplan Meier survival curves for cough, dyspnea, chest pain, fatigue, brain fog, headache, and anosmia/ageusia separately (Appendix S2). Similarly, the risk of any persisting symptom in the unadjusted and adjusted COX models did not significantly differ between the groups (aHR 0.61, 95% CI 0.33-1.15; Table 3). Hazard ratios for individual symptoms were not significant either (Table 3). We found no evidence of effect modification between the SARS-CoV-2 serology status and diabetes, chronic pulmonary disease and cardiovascular disease.

**Figure 3.**
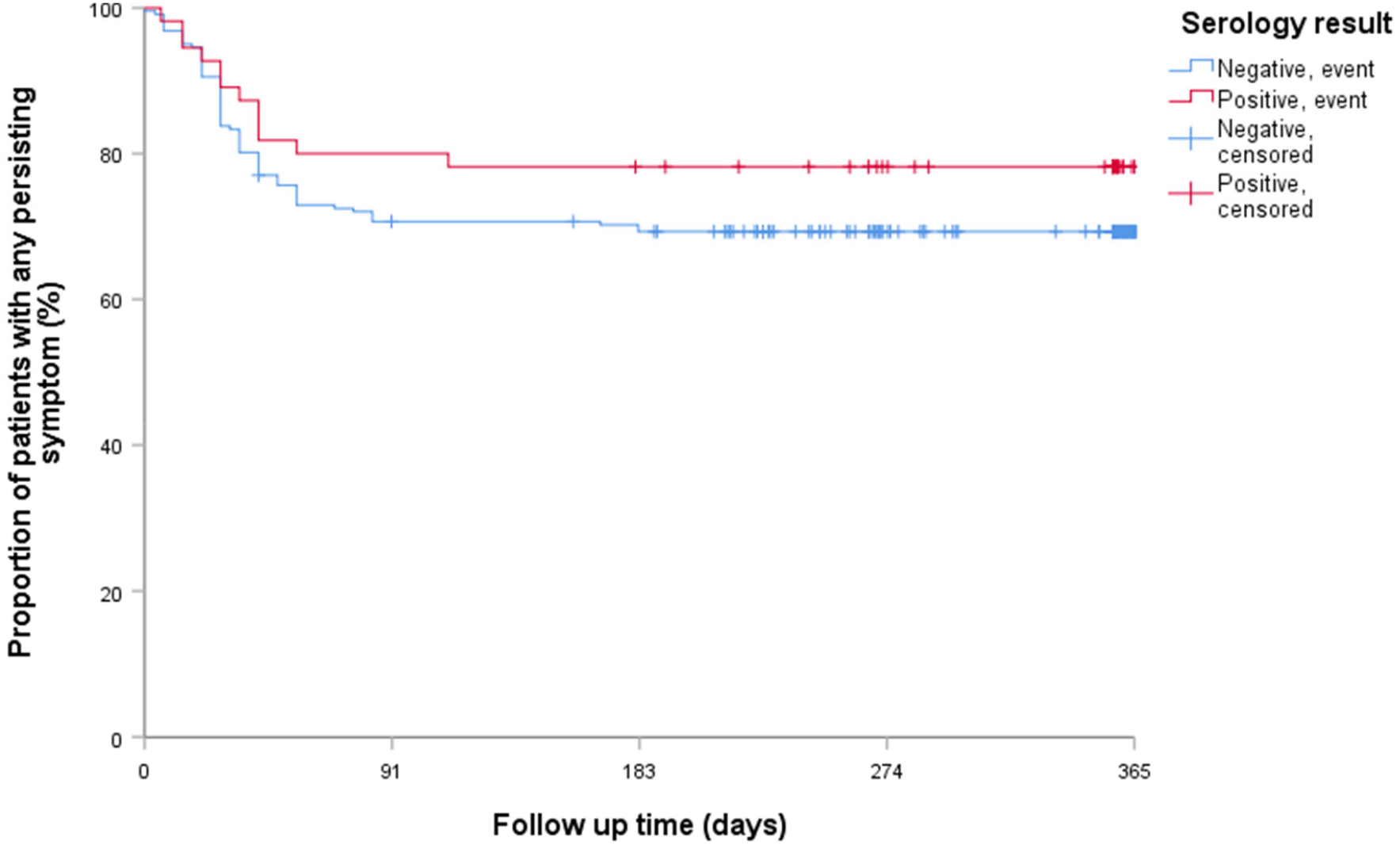
Kaplan Meier curve of the proportion of patients with any persisting symptom over the 12 month follow up

**Table 3.** Adjusted hazard ratio for persisting symptoms until 12 months follow-up for SARS-CoV-2 serology positive and negative participants

| Symptom | Adjusted hazard ratio | 95% CI |
| --- | --- | --- |
| Dyspnea | 0.95 | 0.64-1.42 |
| Cough | 0.92 | 0.65-1.30 |
| Fatigue | 1.01 | 0.66-1.55 |
| Brainfog | 0.85 | 0.59-1.21 |
| Anosmia/ageusia | 0.78 | 0.56-1.09 |
| Headache | 1.01 | 0.73-1.41 |
| Chest pain | 0.96 | 0.69-1.34 |
| Any symptom | 0.61 | 0.33-1.15 |

Also in the sensitivity analyses, 1) censoring patients from the moment they were aware of their COVID-19 status and 2) excluding patients with chronic pulmonary disease and 3) assuming symptom resolution in all participants with a Likert scale symptom score ≤2, no significant differences were found between the two groups in the Cox proportional hazard models.

## Discussion

This is the first blinded study showing that in the 12 months following their moderately severe LRTI, primary care patients with and without confirmed SARS-CoV-2 infection had a comparable HRQoL profile. Albeit a considerable proportion of patients reported persistent symptoms, we found no evidence of a difference in resolution of symptoms over time between patients with and without confirmed SARS-CoV-2 infection.

Thus far, few methodologically rigorous studies have been conducted to determine the long-term sequelae of COVID-19 in primary care patients. In a prospective cohort study of 8786 adults tested for SARS-CoV-2 with RT-PCR (794 positive and 7229 negative) in Norway, 36% of SARS-CoV-2 positive participants reported a “somewhat” or “much” worse health than one year ago at 3-8 months follow-up compared to 18% of RT-PCR negative participants (10). Of all self-reported symptoms over time, only changes in senses of smell and taste and “other symptoms” were reported significantly more frequently at 3-8 months follow-up in RT-PCR positive participants. In a study including 1395 healthcare professionals in Sweden SARS-CoV-2 serology was performed every 4 months (19). Of those 1395 particpants, 323 had had mild or severe symptoms and had positive serology test result and 1072 had negative serology test results and not necessarily had symptoms. At 8 months follow-up, persistent symptoms were more frequently reported in SARS-CoV-2 serology positive than negative participants.

A study performed in Italy on 150 healthcare workers of whom 120 had had COVID-19 (confirmed by RT-PCR) and 30 controls who had not been previously affected by COVID-19 found that neurological deficits and cognitive impairment occurrence at 4 months did not significantly differ between healthcare workers with and without RT-PCR confirmed SARS-CoV-2 infection (20).

In all three studies, follow-up was considerably shorter than in our study and patients were aware of their SARS-CoV-2 status. Besides, controls not necessarily had experienced a respiratory tract infection.

In a cross-sectional analysis of a population-based cohort study in France persisting symptoms at 10-12 months after the COVID-19 pandemic first wave occurred more frequently in patients with self-reported COVID-19 than those without (9). Interestingly however, the differences diminished, except for anosmia, when comparing SARS-CoV-2 serology positive with negative participants. This suggests that the belief in having been infected with SARS-CoV-2 may influence the occurrence of persistent symptoms following COVID-19. This could explain why we found a comparable HRQoL profile between SARS-CoV-2 serology positive and negative participants during the 12 months following a moderately severe LRTI. HRQoL estimates at baseline were comparable to those from a healthy open population (21). It is however important to note that a substantial part of both SARS-CoV-2 serology positive as well as negative participants reported persistent symptoms. This suggests that a moderately severe LRTI can lead to long-term sequelae, irrespective of the causative pathogen.

To our knowledge, this is the first study in which both participants, as well as healthcare professionals and study personnel were blinded for the fact whether study participants had had a SARS-CoV-2 infection or not. This is important as blinding prevents different sorts of bias, like information and recall bias. Another strength of this study is the prospective design of our study and the inclusion of a SARS-CoV-2 serology negative control group of patients with a moderately severe LRTI and the standardized and detailed collection of important prognostic factors (confounders). This allowed us to determine the impact of LRTI etiology on patient’s HRQoL and persistence of symptoms.

Before interpreting our results however, also some possible limitations should be considered. First, misclassification of SARS-CoV-2 serology status due to waning antibodies over time cannot be excluded. However, IgG does not intersect the threshold for seropositivity after 7 months in around 92% of patients and probably in even a larger proportion of patients with more severe symptoms (Den Hartog G, CID 2021). Besides, a previous study in which initial SARS-CoV-2 infection was ascertained, higher antibody titers were associated with more persistent symptoms, making misclassification in COVID-19 patients with persisting symptoms even less likely (7). Therefore, we believe that this did not influence our findings substantially.

Also, it is possible that some patients acquired an (asymptomatic) SARS-CoV-2 infection after the index consultation which may have diluted the contrast between the groups. As patients were asked in each questionnaire whether they experienced SARS-CoV-2 infections after the index consultation, most infections will be registered. Still, asymptomatic infections could have gone undetected. An open population study in the same time period, however, showed a point-prevalence of asymptomatic COVID-19 patients in outpatients of only around 3% (22). Second, participants provided data about the index consultation which was on average seven months ago, and therefore registration of HRQoL and symptoms at that moment may have suffered from inaccurate recollection by study participants. However, HRQoL modelling was mostly performed on prospective data as only the SF-36 scores from 2 weeks prior to the index consultation was determined retrospectively.

Furthermore, it is unlikely that inaccurate recall differed substantially between SARS-CoV-2 serology positive and negative participants due to blinding. Finally, our study population was not yet vaccinated against COVID-19 and probably not previously infected by SARS-CoV-2 at the time of the index consultation. It remains unclear whether presence of SARS-CoV-2 antibodies, through vaccination of acquired infection would lead to fewer persisting symptoms.

Our results indicate that persisting symptoms occur in a substantial proportion of patients following a LRTI. Observed long-term sequelae after moderately severe SARS-CoV-2 LRTI in primary care might therefore not be considered as a SARS-CoV-2 specific phenomenon. Further studies looking into the persistence of symptoms after LRTI, comparing different pathogens are therefore warranted.

## Conclusion

In conclusion, we saw no clear differences in HRQoL, nor in the duration of symptoms between primary care treated patients with and without confirmed SARS-CoV-2 infection, but all suffering a moderately severe LRTI, in the first year after infection. However, a substantial proportion of patients who had a moderately severe LRTI did have symptoms beyond 4 weeks. Our results suggest that persisting symptoms might not be considered specific for SARS-CoV-2, but can also occur in LRTI’s with another etiology. Further research regarding persisting complaints after LRTI should therefore not only focus on COVID-19 infections, but also on respiratory infections caused by other pathogens.

## Data Availability

All data produced in the present study are available upon reasonable request to the authors

---

**Appendix S1.**
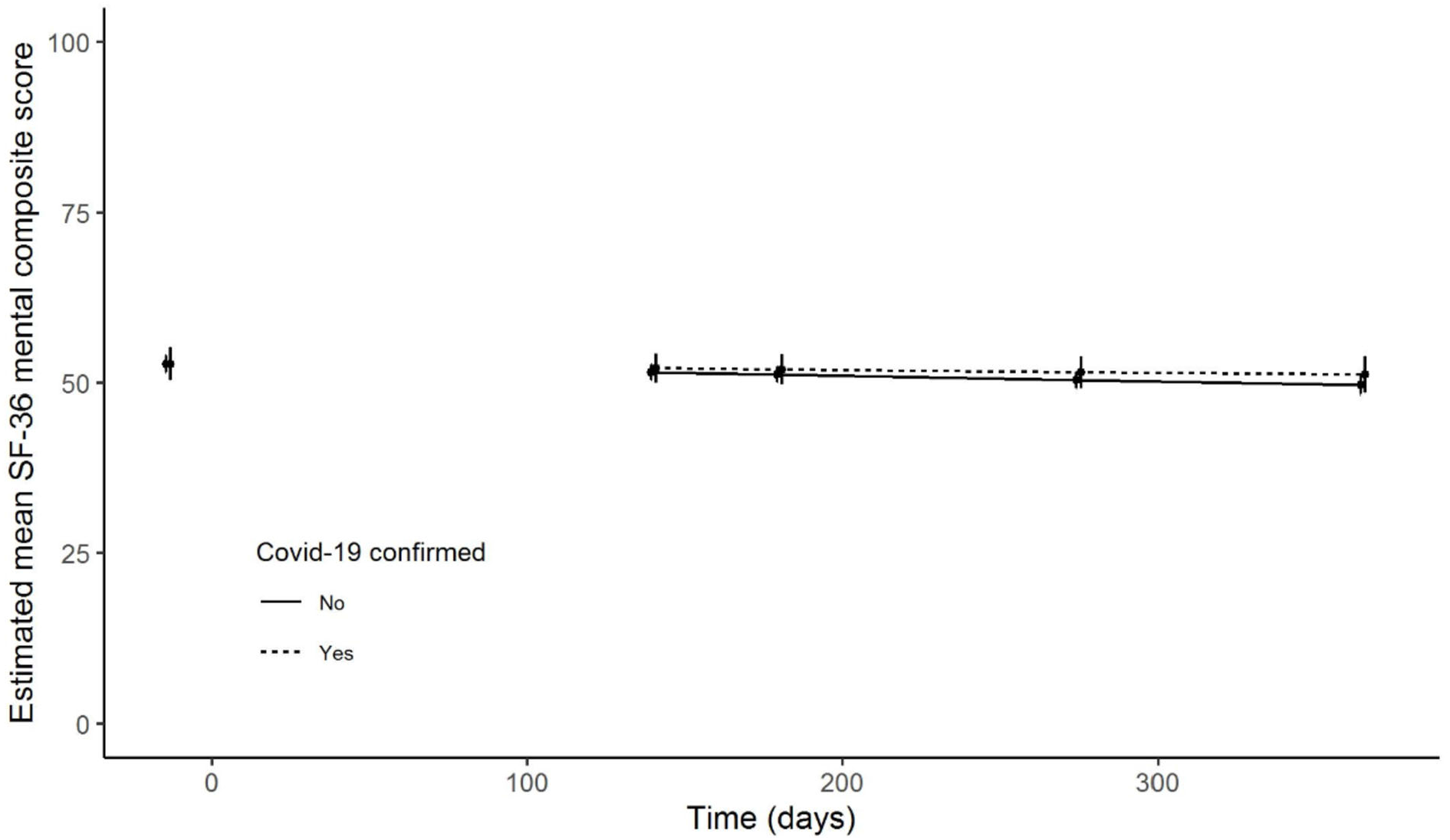
Figure appendix S1 Estimated mean MCS score (SF-36) over time, based on mixed model.

**Appendix S2.**
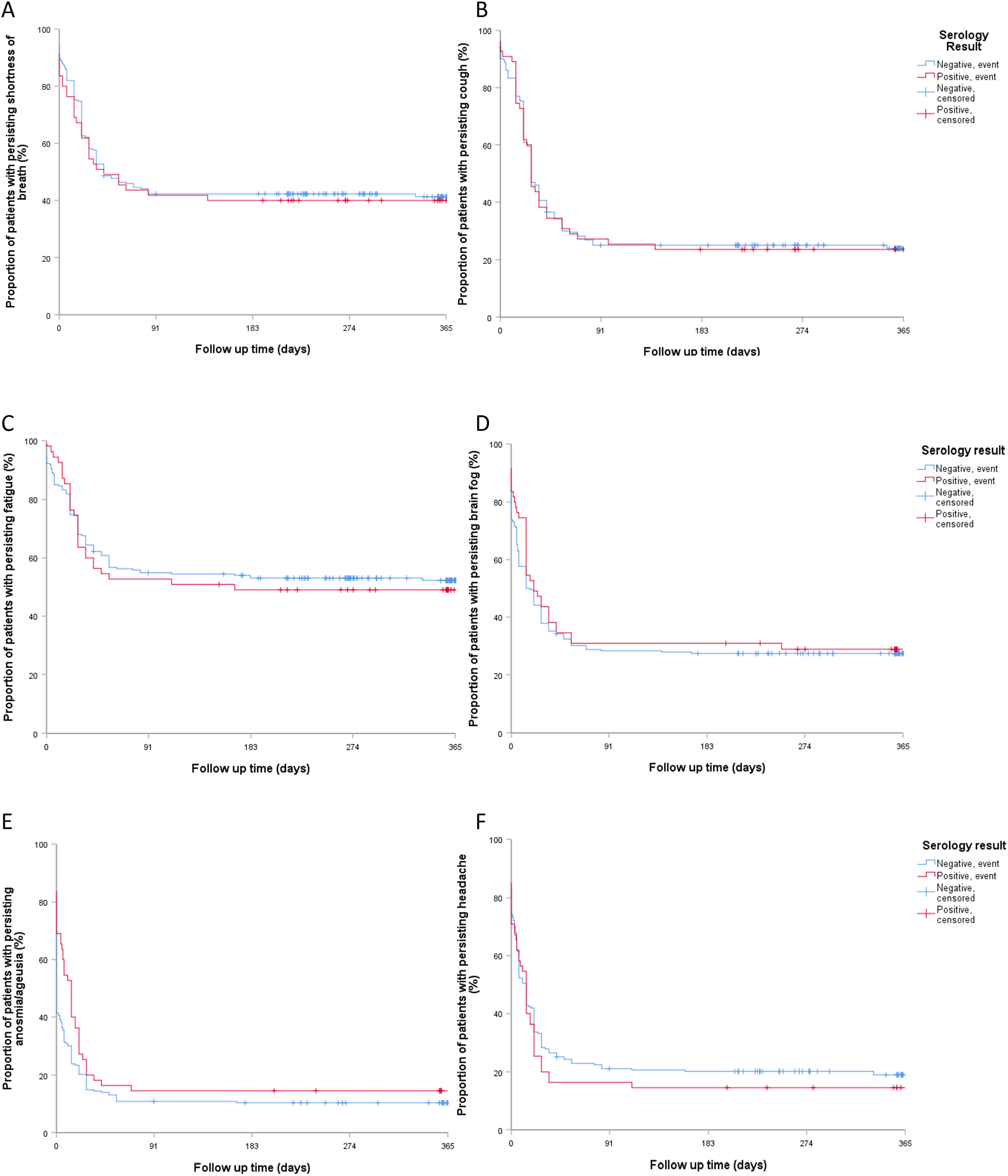
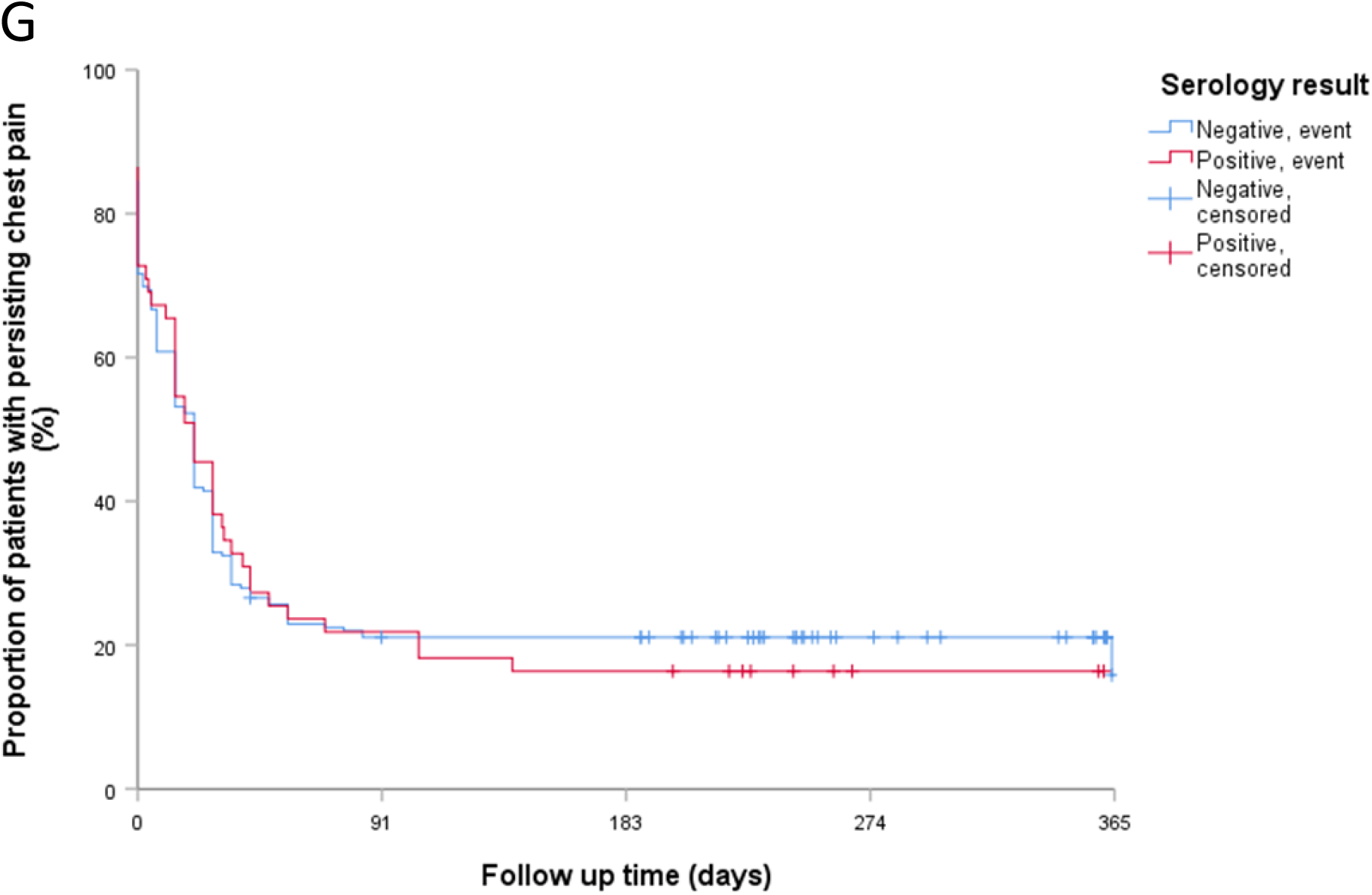
Appendix S2 Kaplan Meier survival curves of proportion of patients with persisting (A) shortness of breath, (B) cough, (C) fatigue, (D) brain fog, (E) anosmia/ageusia, (F) headache and (G) chest pain over a 12 month follow up

